# Optimization of an adult immunization program in Canada

**DOI:** 10.1101/2024.06.10.24308663

**Authors:** Nathan Locke, Mike Paulden, Shannon E. MacDonald, Stephanie Montesanti, Ashleigh Tuite, Karsten Hempel, Wade McDonald, Ellen Rafferty

## Abstract

**Background:** Provincial decisions to fund a new immunization program are generally made on a case-by-case basis, without systematic consideration of how the new immunization program may fit within the larger provincial immunization portfolio.

**Aim:** The goal of this study was to develop evidence and tools to guide policy-makers in making fiscally and ethically responsible decisions on which adult immunization programs to include in their portfolio under various constrained budgetary scenarios.

**Methods:** Using previously published infectious disease models, cost-utility data was estimated for adult pneumococcal, influenza, pertussis, and shingles immunization programs. This data was then inputted into a newly developed constrained optimization model to determine portfolios of immunization programs that maximize either population health or incremental net monetary benefit, subject to a budget constraint. Sensitivity analyses were conducted on model parameters such as vaccine costs, cost-effectiveness thresholds, and the budget constraint.

**Results:** Optimized solutions changed dramatically based on the number of immunization programs included, total budget, what was optimized for (i.e., population health or incremental net monetary benefit), the cost-effectiveness threshold and the assumed vaccine prices. Maximal health gains and budget spending was achieved when optimizing based on population health. Reductions in health gains and budget spending were observed at a CAN$50,000 cost-effectiveness threshold, and at a CAN$30,000 threshold, the budget was significantly underutilized and health gains were noticeably reduced.

**Conclusion:** If budgets for the adult immunization portfolio are fixed, then shifting to more expensive programs that offer large health benefits may be preferable. However, if budgets can be spread across various public health programs (i.e., childhood immunization, well-baby programs), it may make more sense to optimize based on cost-effectiveness. Constrained optimization tools could improve goals-based decision-making and allow for transparent and effective methods to make allocation decisions.

**Highlights:**

- Optimized solutions changed dramatically based on the number of immunization programs available, total budget, and the cost-effectiveness threshold.
- If budgets for the adult immunization portfolio are fixed, then shifting to more expensive programs that offer large health benefits may be preferable.
- If budgets can be spread across various public health programs, it may make more sense to optimize based on cost-effectiveness.
- Constrained optimization tools could improve goals-based decision-making and allow for transparent and effective methods to make allocation decisions.

## I. Introduction

Healthcare systems across the globe are under increasing financial strain. Canada has one of the highest health care spending to gross domestic product (GDP) ratios (12.3%)(1) amongst Organisation for Economic Co-operation and Development countries, behind only the United States, Germany and the United Kingdom (UK). This ratio has been steadily increasing since 1975, when healthcare made up 7% of total GDP spending in Canada(2). By taking up a larger percentage of total spending, the healthcare system puts pressure on other sectors of the economy, making it essential to identify ways to increase the efficiency of spending in healthcare.

Evidence suggests that preventive and public health interventions are generally good value for the money; and cuts to public health budgets could potentially increase costs in acute care medicine and the wider economy(3). A study from the UK found that additional spending in public health produces more health per dollar spent than identical spending in treatment activities(4). However, public health typically constitutes a substantially smaller percentage of the overall healthcare budget (6.1% of total health spending in Canada)(1), leading to public health and immunization departments grappling with tough decisions about which programs to fund to optimize health and achieve the best use of scarce financial resources.

Across Canada, one of the main responsibilities of provincial/territorial public health departments is immunization. Since immunization programs make up a substantial proportion of public health budgets, it is important that policy-makers understand the benefits and costs of these programs, and make decisions about which large-scale immunization programs they should fund. While historically immunization programs have been great value for the money(5), new programs, especially adult immunization programs, can be very expensive, raising questions about their cost-effectiveness. With multiple new adult vaccines either recently licensed or coming down the pipeline, including vaccines for influenza(6), respiratory syncytial virus (RSV)(7), and Lyme disease(8), among others, public health decision-makers across the world will need to make budgetary decisions, including whether to expand their budgets, replace pre-existing adult immunization programs, or exclude any new vaccines.

Public health researchers and policy-makers need evidence-based research and methods for determining where the limited amount of immunization budget available to them is most efficiently spent. The objectives of this research were to: 1) conduct a case study on the optimal mix of adult immunization programs in a provincial public health system based on their incremental net monetary benefit (INMB) or health benefits (quality adjusted life years – QALYs); 2) develop an interactive optimization model that can be adapted by various jurisdictions. This analysis can provide insight into the trade-offs associated with budgetary changes to a portfolio of adult immunization programs, as well as inform decisions around where new adult vaccines may fit, both compared to other programs for that same disease, and within the larger immunization portfolio.

## II. Methods

In this analysis, we conducted a constrained optimization experiment to determine the optimal (i.e., most efficient) mix of adult immunization programs that maximized either health outcomes or INMB under specific budgetary constraints. We took the perspective of a regional public health program, with the province of Alberta, Canada as the case study. We focused on sustained large-scale immunization programs available in Canada that targeted either the entire adult population or a large subset of the population (e.g., specific age groups) on an ongoing or recurring (i.e., annual/seasonal) basis. We selected vaccine-preventable diseases, with specific immunization programs for adults that all or some Canadian provinces had publicly funded, including influenza, herpes zoster (shingles), pertussis in pregnant women and pneumococcal disease. We did not include vaccines that are currently funded nationally in Canada (e.g., COVID-19), or were not licensed at the time of the analysis.

This analysis involved four key steps: 1) identification of peer-reviewed infectious disease models and relevant immunization scenarios; 2) estimation of health outcomes for each immunization scenario using the identified infectious disease models; 3) application of healthcare costs and QALYs to the estimated health outcomes and calculation of the INMB; and 4) optimization of the adult immunization portfolio based on INMB or health benefit, and subject to a budget constraint.

We received ethical approval from the University of Alberta Health Research Ethics Board (Ethics ID: Pro00112164) and funding from the Canadian Institute for Health Research (CIHR) Catalyst Grant: Impacts of financial and organizational restructuring of public health (Grant Number: 435188). CIHR had no role in the design or conduct of the study.

### Identification of peer reviewed disease models & immunization scenarios

To estimate health outcomes for each disease, we first identified pre-existing infectious disease models that were suitable for the needs of this analysis. We conducted a rapid review of the literature to identify relevant infectious disease models. Inclusion criteria for the disease models included: 1) full text article available; 2) English language; 3) evaluated an immunization program; 4) Canadian setting; and 5) covered one of the four diseases under study (i.e., pertussis, shingles, pneumococcal, and influenza). We identified 64 articles that described potentially relevant infectious disease models. We then selected the four models (9-12) used in the analysis based on the following criteria: 1) model was available and adaptable; 2) Canada-specific (with preference given to those that were Alberta-focused); 3) included the vaccines of interest; 4) time horizon of the analysis; 5) year of study; 6) stratification of the results (e.g., age groups, sex, health risk factors); and 7) health outcomes captured.

Table 1 lists the immunization scenarios that we included in the analysis. These scenarios were selected based on the capabilities of the infectious disease models available for use in this analysis and expert opinion on which vaccines would be considered for inclusion in their adult immunization portfolio. Multiple scenarios are included to capture the effects on health outcomes that occur from changes in infectious disease models parameters such as coverage rate, the age group eligible for vaccination, or the type of vaccine used. A baseline scenario is included in each disease’s set of scenarios, where no adult immunization program is offered (except for influenza, where it is assumed those under 20 are vaccinated at the rates reported in Appendix Table A.1). The baseline scenarios allow us to compare the performance of the adult immunization programs individually and against each other. Table 2 provides information on the effectiveness and costs of the vaccines listed in Table 1.

**Table 1.**
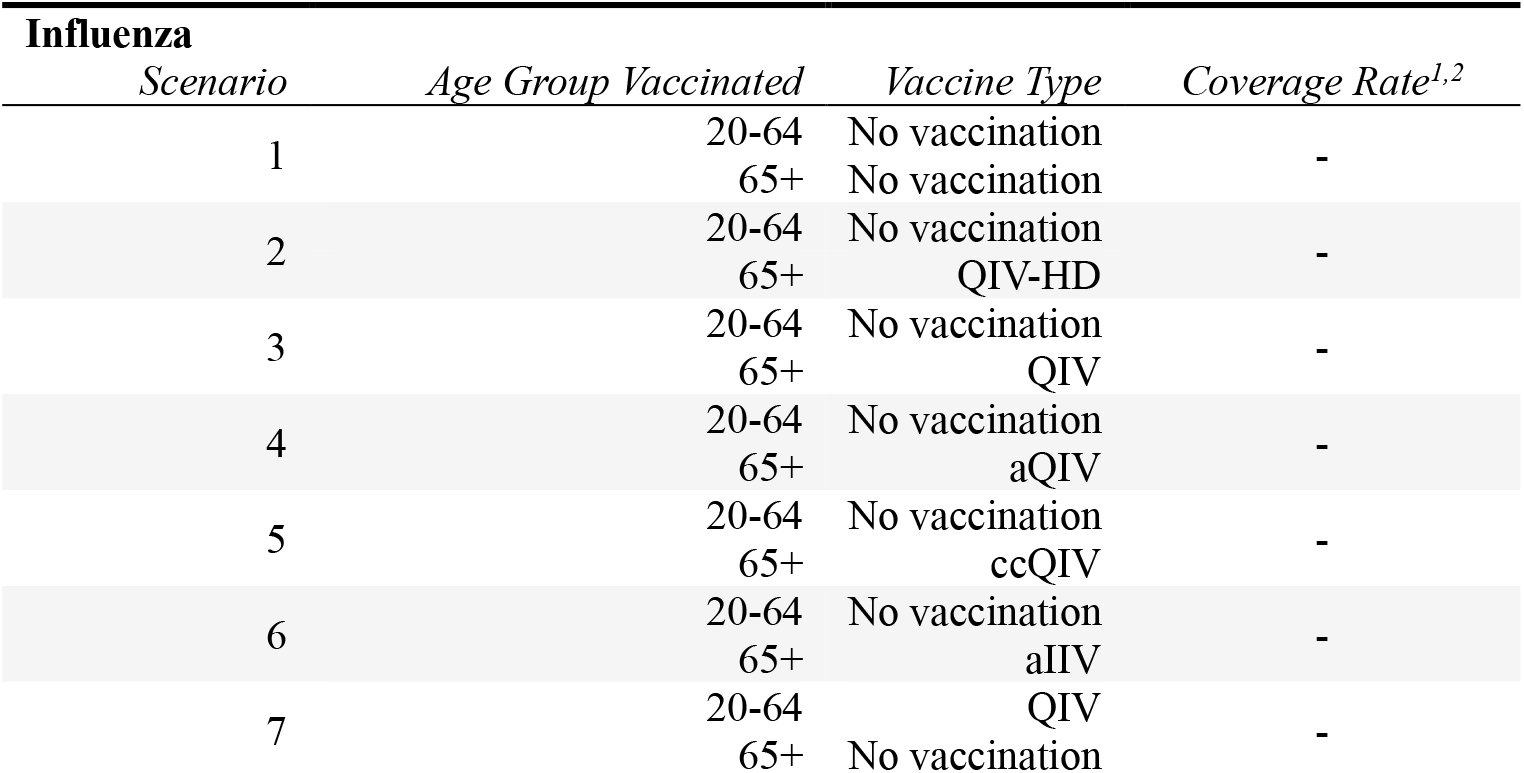
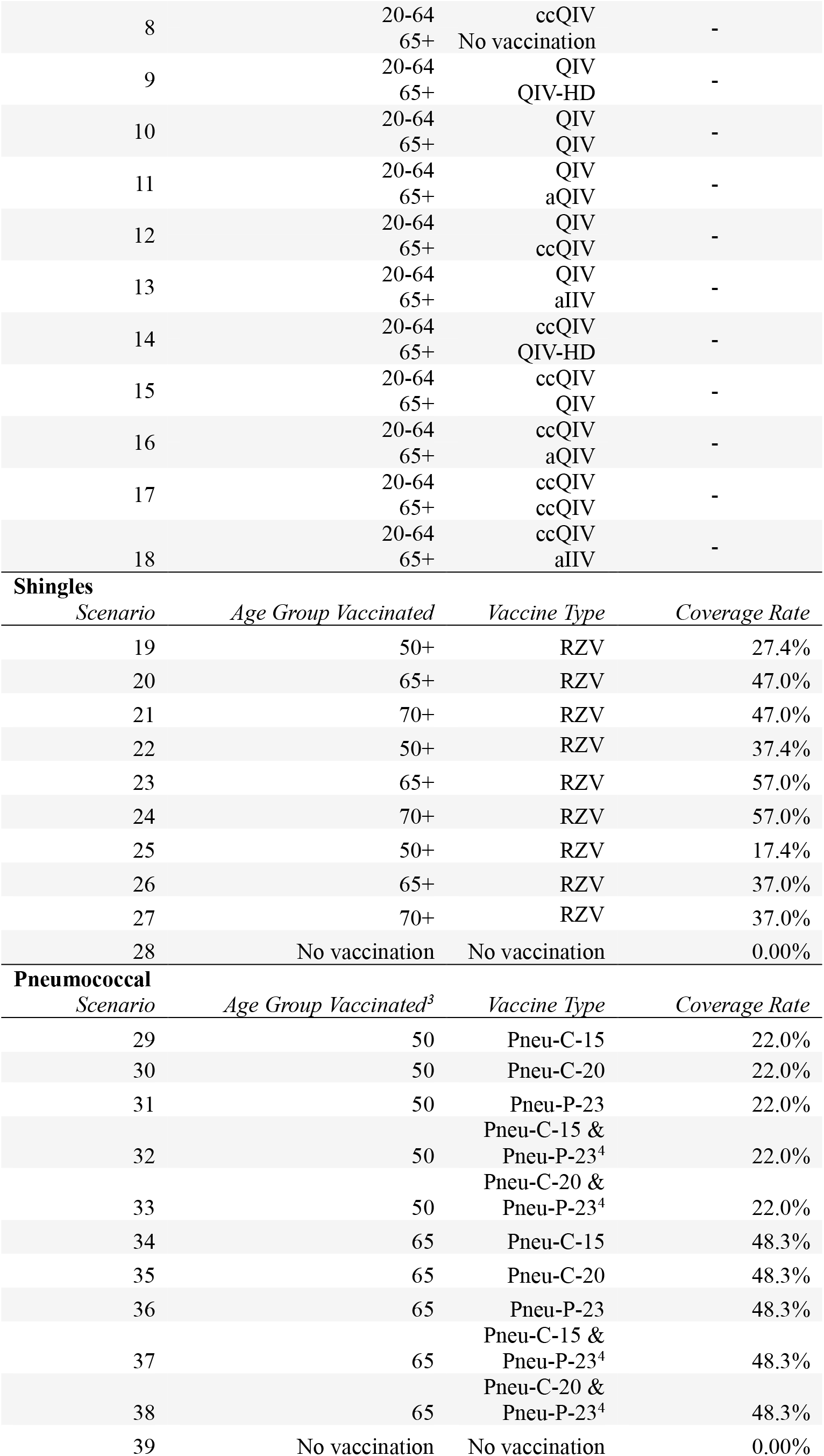
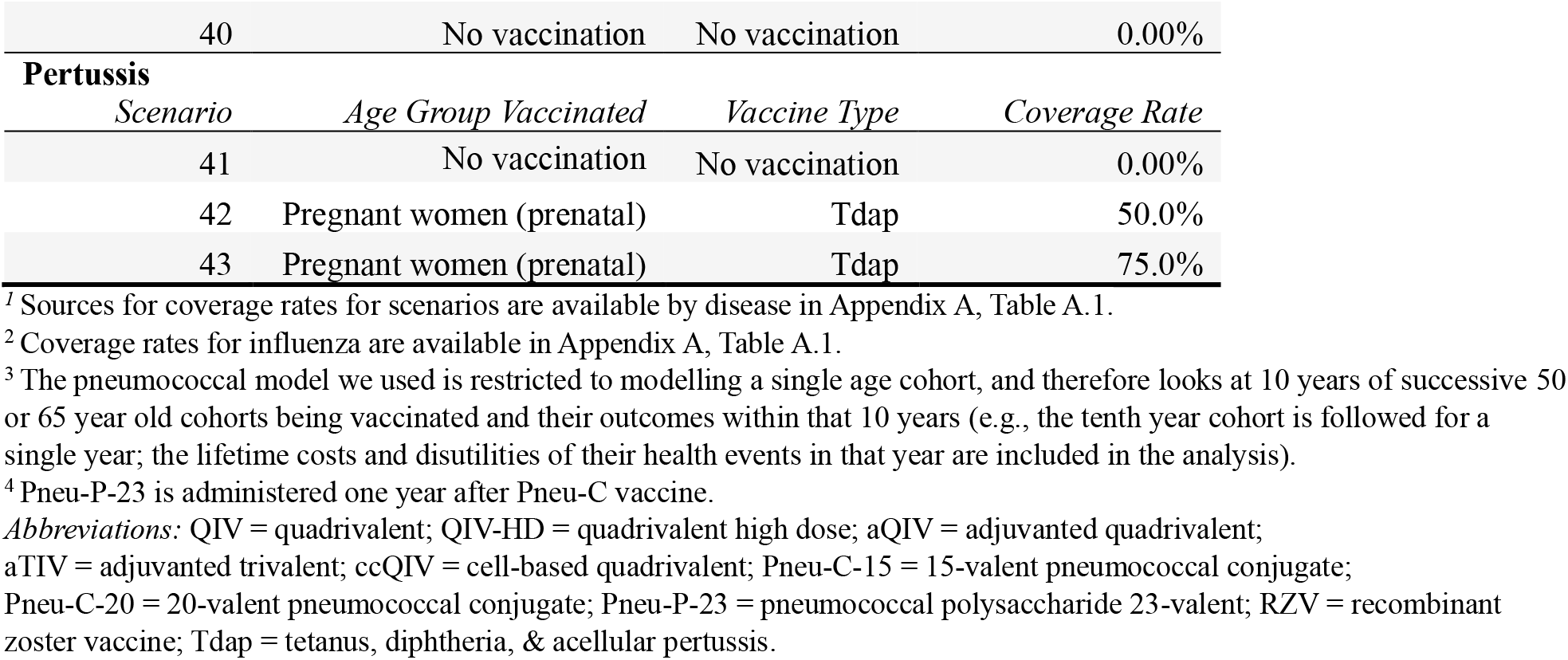
Immunization Scenarios, by Disease and Vaccine Characteristics.

**Table 2.** Vaccine Effectiveness and Costs.

| *Vaccine Type* | *aVE (65+ yrs)* | *aVE (20-64 yrs)* | *aVE (6-19 yrs)* | *aVE (<6 yrs)* | *Cost (per dose)<sup>2,3</sup>* |
| --- | --- | --- | --- | --- | --- |
| **Influenza** |  |  |  |  |  |
| QIV | 0.383 | 0.510 | 0.554 | 0.554 | \$44 |
| QIV-HD | 0.511 | n/a | n/a | n/a | \$76 |
| aQIV | 0.531 | n/a | n/a | n/a | \$72 |
| aTIV | 0.430 | n/a | n/a | 0.860 | \$70 |
| ccQIV | 0.401 | 0.534 | 0.546 | 0.546 | \$49 |
| **Pneumococcal** |  |  |  |  |  |
|  | *aVE VT-IPD* | *aVE ST3-IPD* | *aVE VT-CAP* | *aVE ST3-CAP* |  |
| Pneu-C-15 | 0.600 | 0.260 | 0.450 | 0.156 | \$69 |
| Pneu-C-20 | 0.600 | 0.260 | 0.450 | 0.156 | \$79 |
| Pneu-P-23 | 0.470 | 0.020 | 0.200 | 0.020 | \$37 |
| **Shingles** |  |  |  |  |  |
|  | *aVE (50-69 yrs)* | *aVE (70+ yrs)* |  |  |  |
| RZV | 0.970 | 0.910 | | | \$104<sup>4</sup> |
| **Pertussis** |  |  |  |  |  |
|  | *aVE* |  |  |  |  |
| Maternal Tdap | 0.800-0.900 | | | | \$60 |

<sup>1</sup> Sources for vaccine effectiveness are available in Appendix A, Tables A.2, A.3, A.4, A.5, and A.6.
<sup>2</sup> The vaccine prices presented here for influenza, shingles and pertussis assume a 50% reduction in the US list prices for the vaccines (vaccine prices for pneumococcal determined from Canadian sources). All vaccines also assume an administration cost of \$21.
<sup>3</sup> Sources for vaccine costs are available in Appendix A, Table A.7
<sup>4</sup> Two doses are administered as per recommendations(13).
*Abbreviations:* aVE = absolute vaccine effectiveness; VT = vaccine-type; ST3 = serotype 3; IPD = invasive pneumococcal disease; CAP = community-acquired pneumonia; QIV = quadrivalent; QIV-HD = quadrivalent high dose; aQIV = adjuvanted quadrivalent; aTIV = adjuvanted trivalent; ccQIV = cell-based quadrivalent; Pneu-C-15 = 15-valent pneumococcal conjugate vaccine; Pneu-C-20 = 20-valent pneumococcal conjugate vaccine; Pneu-C-23 = pneumococcal polysaccharide 23-valent vaccine; RZV = recombinant zoster vaccine; Tdap = tetanus, diphtheria, & acellular pertussis.

### Health Outcome Data

We used the four identified infectious disease models to estimate number of vaccines provided and health outcomes (e.g., infections, physician visits, hospitalizations, number of vaccinations) for each immunization scenario. Each model was run over a 10-year time horizon for consistency between scenarios. All models were adjusted to represent the Alberta population in terms of size and age-groups.

The model selected for influenza is an age-structured stochastic compartmental model with a 10-year time horizon(11). The outcomes we estimated using this model included annual vaccinations, influenza symptomatic infections, physician visits, emergency department visits, hospitalizations, ICU cases, and deaths. Estimates of these health outcomes are broken down by year and into three age groups: children (<20), adults (20-64), and seniors (65+). The pneumococcal model is a Markov Chain model developed by the National Advisory Committee on Immunization for the evaluation of newly licensed pneumococcal vaccines(10). The model outputs the number of cases of invasive pneumococcal disease (IPD), community acquired pneumonia (inpatient or outpatient severity separately), post-meningitis sequelae (mild or severe separately) and deaths. Our analysis looks at two age cohorts, where vaccination was provided at age 50 or 65. For pertussis, we used an agent-based model developed by Hempel et al. (2023)(12) that included maternal vaccination against pertussis. The model is a stochastic dynamic infectious disease model that estimates pertussis cases across 1-year age increments between 0 and 99.

We selected an agent-based model by Rafferty et al. (2018)(9) to estimate shingles-related health outcomes associated with immunization scenarios. The outcomes produced by this model include vaccinations, infections, cases of PHN, and hospitalizations.

### Calculating Total Costs, QALYs and INMB

Once we had produced health outcomes for each vaccine scenario, we then applied healthcare costs and QALY estimates to these outputs., We used costs and QALY estimates for influenza from Fisman et al. (2011)(14), for pneumococcal from PHAC (2023)(10), for pertussis from McGirr et al. (2019)(15) and for shingles from Rafferty et al. (2021)(16) and Friesen et al. (2017)(17). All costs were adjusted to 2022 Canadian Dollars and both costs and QALYs were discounted at a rate of 1.5% per year.

Using the cost and benefit outputs, we estimated INMB for each immunization scenario in comparison to baseline (no adult vaccination). We calculated INMB using both a CAN$30,000 and CAN$50,000 threshold (18, 19).

### Optimization

Our optimization analysis was conducted in accordance with good practice recommendations set out by the Professional Society for Health Economics and Outcomes Research (Crown et al., 2017; Crown et al., 2018). Two different optimization approaches were used in this analysis, varying only in how their objective function is defined. One approach is concerned with maximizing INMB, the other is concerned with maximizing population health based on QALYs (i.e., immunization scenarios are compared based on effectiveness rather than cost-effectiveness). Occurring under a budget constraint, our approach sought to maximize the total INMB or health benefits achieved in our selected combination of immunization program scenarios. Only one immunization scenario can be selected for each disease and scenarios must be fully funded, so our optimization approach is based on integer linear programming. The notation for the objective function and constraints are as follows in Equation Set A:

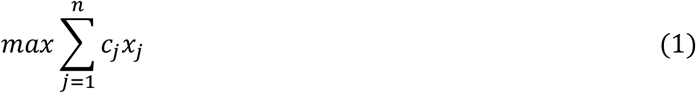

subject to:

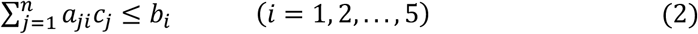

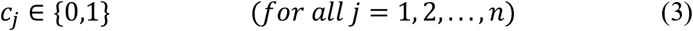

The objective function (line 1) aims to maximize the sum of benefits from a limited combination of immunization scenarios, where *j* represents a given immunization program scenario, *x* represents the benefit of immunization program scenario *j*, and *c* represents our choice variable (either a 1 or 0) on whether a given scenario is included in the portfolio. The objective function is subject to four constraints: an integer constraint on the choice variable (line 3), a budget constraint (line 2), and a disease constraint which limits the number of immunization programs that can be selected per disease in the optimal solution to one (line 2). This final constraint on programs per disease is included as our immunization program scenarios are mutually exclusive within each disease category and we did not want to include two scenarios that only differ in their coverage rate or age (for pertussis, shingles, pneumococcal), or vaccinate a specific age group with two types of vaccines (for influenza, pneumococcal). Line 2 includes *a*_*ij*_, where *a*_1*j*_ represents the cost of vaccination program scenario *j*, and *a*_2*j*_ to *a*_5*j*_ which indicate what disease scenario *j* targets. The sum across each *i* must be less than or equal to its corresponding constraint value of *b*_*i*_ which is a vector of upper bound constraints with a length of five: with *b*_1_ representing the adult immunization budget and the remaining four constraints, which limit the number of immunization programs per disease, having a value of one.

In the main scenario analysis, we estimated both health-maximizing and INMB at a hypothetical CAN$500M adult immunization budget. We estimated these results for portfolios that contain one, two, three or four immunization program scenarios and at two cost-effectiveness thresholds: CAN$30,000/QALY and CAN$50,000/QALY. With the exception of pneumococcal vaccines, vaccine costs used in the main scenario represent a 50% reduction of published estimates, under an assumption that public pricing is lower than vaccine list prices from the United States, and so that all diseases could be considered in the cost-effectiveness optimization. Pneumococcal vaccine prices were obtained from Canadian sources and therefore did not include this 50% reduction. We then conducted sensitivity analyses around the total adult immunization budget, looking at lower (CAN$100M) and higher (CAN$1B) budgets, higher vaccine costs (i.e., costs consistent with published estimates), and a cost-effectiveness threshold of CAN$100,000/QALY. The selection of the lower budget threshold was based on an assumption that the actual 10-year budget is likely more than CAN$100M(20), whereas the upper bound budget threshold was set at CAN$1B because values beyond this amount did not yield new optimization solutions.

We solved the optimization problem in *R(21)* using the package *FLSSS(22)*. Its performance was validated by comparing its results against the solutions of Microsoft Excel’s Solver(23), which uses an appropriate branch and bound method (24-26). We then input the optimization model into R shiny, which allows users to change key parameters, including vaccine price, cost and quality of life loss associated with health events, the type of analysis (cost-effective vs. health maximizing solutions), the immunization program scenarios, the total budget and the cost-effectiveness threshold. The model is available at https://eshiny.ihe.ca/cvop/.

## III. Results

Overall, we were able to optimize the adult immunization portfolio using both the health maximizing and the INMB approach with thresholds of CAN$30,000 and CAN$50,000, assuming a budget of CAN$500 million. We estimated all results assuming a one, two, three or four immunization program portfolio. The results of this main scenario, including total health benefit (QALYs gained), percentage of the budget used, estimated incremental cost-effectiveness ratio (ICER) based on the portfolio selected, and immunization program(s) selected, at each portfolio size, are presented in Table 3.

**Table 3.**
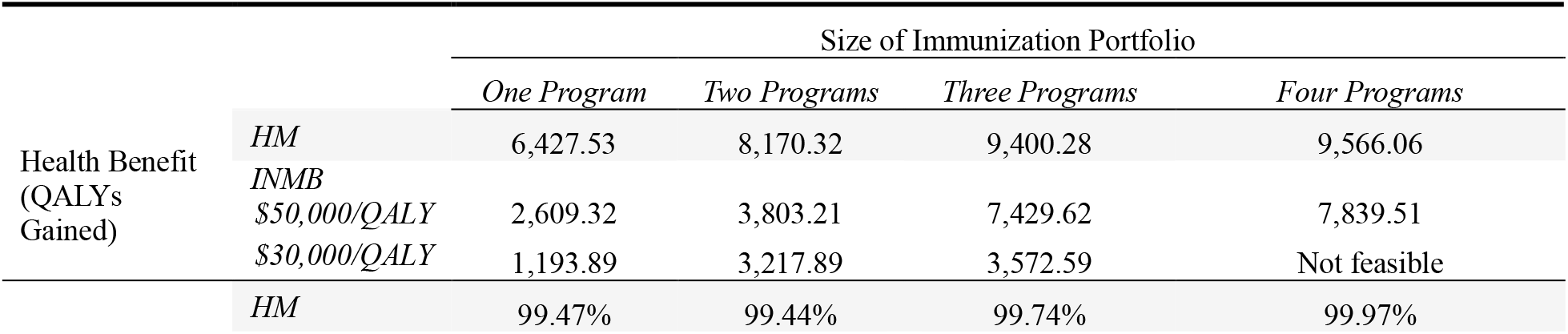
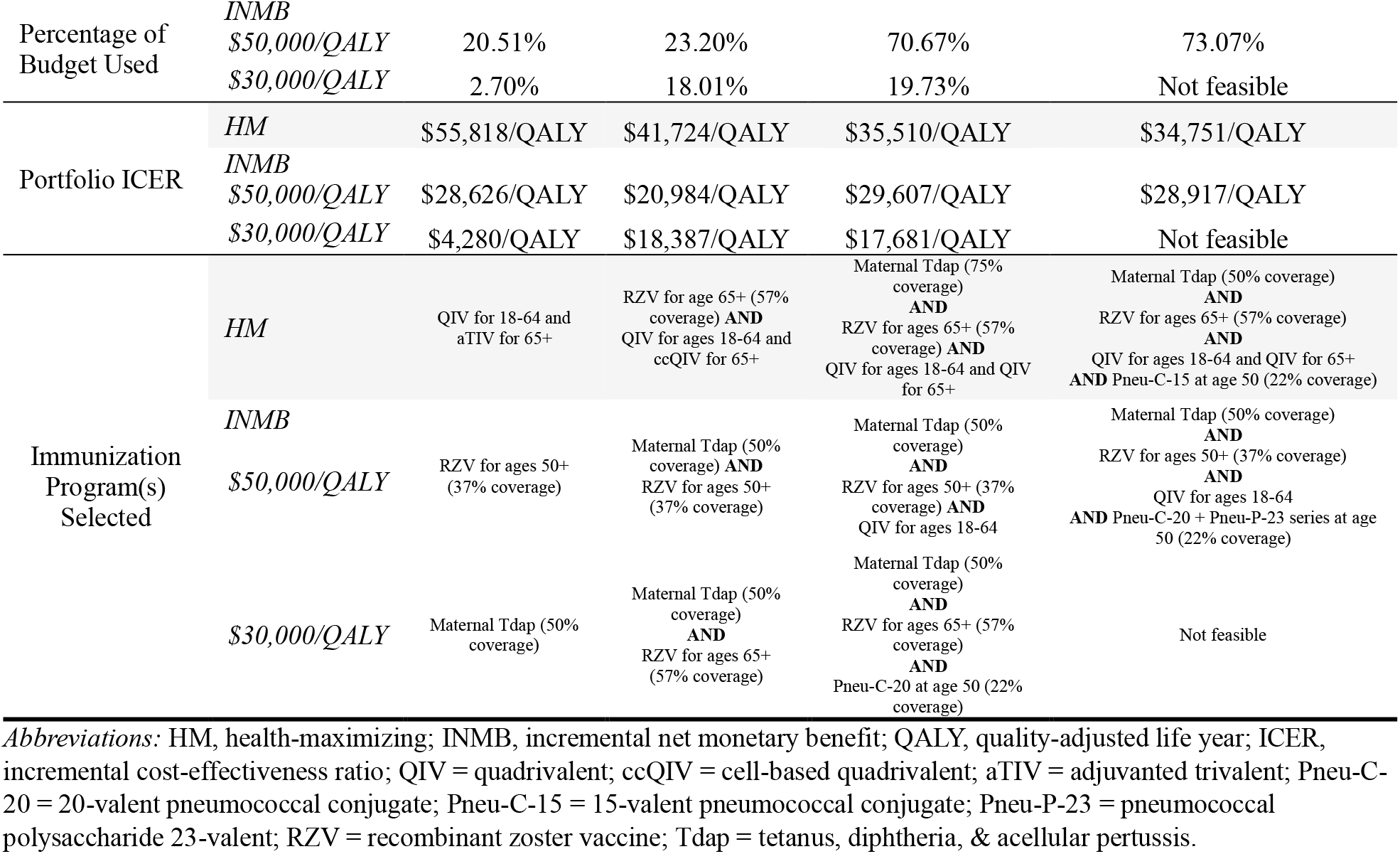
Vaccine Portfolio Selections, Main Scenario, CAN$500M Budget.

Our results found that the health-maximizing approach always achieves a greater health benefit to that of the INMB approach. As influenza immunization scenarios offer the largest net gains in health benefits, the health-maximizing approach selects the influenza scenario that provides the most QALYs possible while remaining under the CAN$500M budget for its one program portfolio. Table 4 provides an overview of the average health benefits and vaccination program costs within each disease category. At a cost-effectiveness threshold of CAN$50,000/QALY, the INMB optimization allocates its budget to a shingles program for the one program portfolio. Note, this optimization did not select the most cost-effective immunization scenario overall, which was the Tdap immunization program (50% coverage). While the pertussis programs are very cost-effective, the total health benefit that they offer is smaller than that of shingles programs.

**Table 4.**
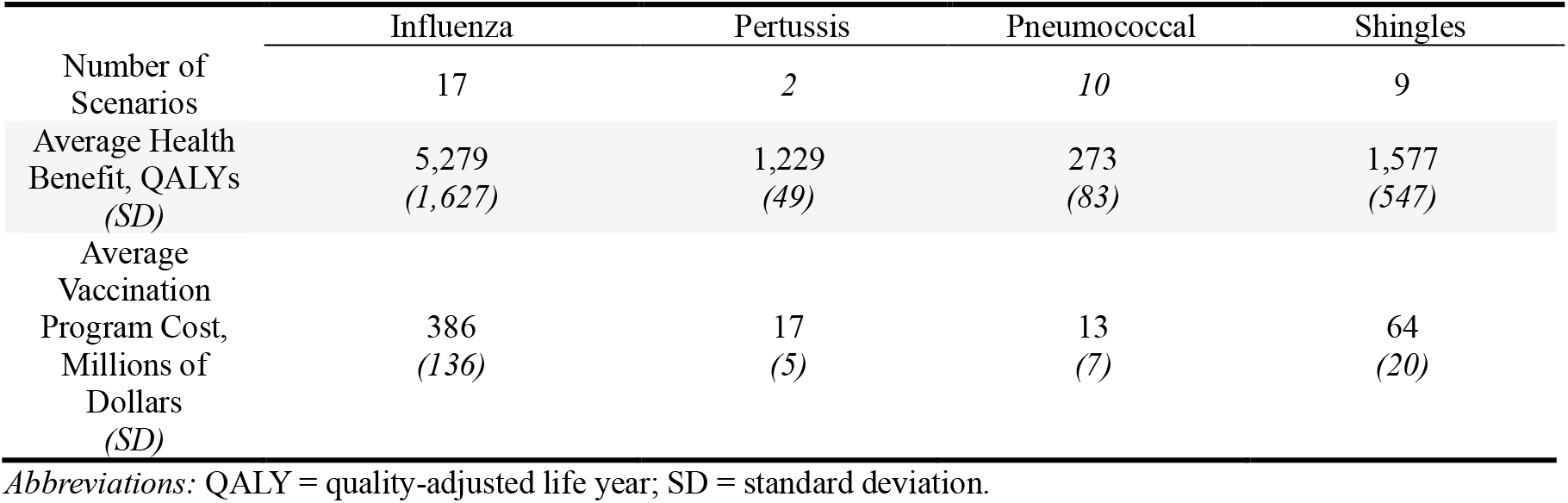
Average Health Benefits and Vaccination Program Costs of Scenarios, by Disease.

|  | Influenza | Pertussis | Pneumococcal | Shingles |
| --- | --- | --- | --- | --- |
| Number of Scenarios | 17 | 2 | 10 | 9 |
| Average Health Benefit, QALYs (SD) | 5,279 (1,627) | 1,229 (49) | 273 (83) | 1,577 (547) |
| Average Vaccination Program Cost, Millions of Dollars (SD) | 386 (136) | 17 (5) | 13 (7) | 64 (20) |

*Abbreviations:* QALY = quality-adjusted life year; SD = standard deviation.

The only portfolio without a solution was the four program portfolio at a threshold of CAN$30,000 (see Table 3). We found no feasible solution for this portfolio because there were no influenza programs deemed cost-effective at that threshold. For this reason, the INMB optimization with a CAN$30,000/QALY threshold must instead select between immunization scenarios for pertussis, pneumococcal, or shingles. As seen in Table 4, shingles would generally offer the most potential health benefit out of these three diseases, yet the optimization begins by selecting a pertussis vaccination scenario instead. Unlike the INMB optimization with a CAN$50,000/QALY threshold, shingles scenarios do not have a sufficiently large enough health benefit over pertussis to be favoured at a cost-effectiveness threshold of CAN$30,000/QALY. To illustrate the trade-off between cost-effectiveness and net health gains, Figure 1 shows how the INMB of pertussis and shingles immunization scenarios change as the cost-effectiveness threshold increases. Shingles immunization scenarios generally achieve a positive INMB at just below CAN$30,000/QALY; however, it is only around the CAN$50,000/QALY cost-effectiveness threshold that one of the shingles scenarios can achieve a larger INMB than a pertussis scenario.

**Figure 1.**
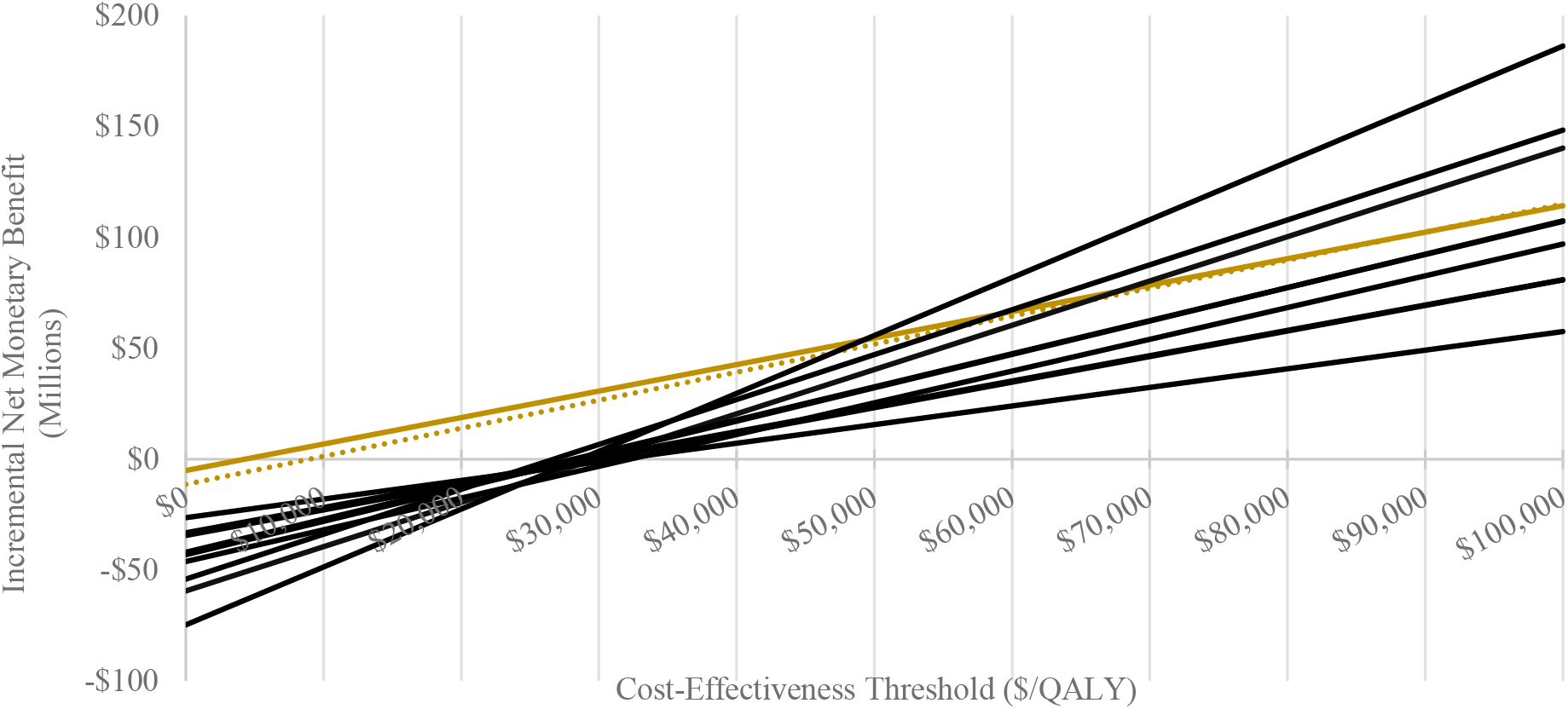
Incremental Net Monetary Benefit of Pertussis (Gold) and Shingles Vaccination (Black) Scenarios, by Cost-Effectiveness Threshold

We also present the calculated ICER of each portfolio solution. The maximum ICER value is CAN$55,818/QALY, occurring under the one program portfolio for the health-maximizing approach. As the size of the health-maximizing portfolio increases, the ICER continuously decreases. The association between portfolio size and the ICER value is a byproduct of the dynamics between scale and cost-effectiveness of immunization scenarios between disease types. Influenza vaccination programs scenarios offer the largest health benefits, whereas pneumococcal vaccination programs offer the smallest amount. At the same time, pneumococcal programs are some of the most cost-effective scenarios and influenza programs are the least. Because of these dynamics, the health-maximizing approach is gradually adding more cost-effective disease categories as portfolio size increases. Moreover, it may need to reallocate budget to move from a one-program portfolio to a two-program one (99.47% of the budget is used for the one-program portfolio), so it must also consider a more cost-effective influenza program, which leads to the ICER value decreasing further.

In addition to our main scenario, we conducted additional sensitivity analyses which involved either using the full cost of each vaccine or a CAN$100,000/QALY cost-effectiveness threshold. Results of these sensitivity analyses are presented in Appendix A (Tables B.1 and B.2). Increasing the cost-effectiveness threshold to CAN$100,000/QALY allows additional influenza scenarios with an ICER of more than CAN$50,000/QALY to be considered in the optimization.

Lastly, we also conducted a sensitivity analysis around the total budgetary constraints, where the health-maximizing optimization and CAN$50,000/QALY INMB optimization for the main scenario were re-estimated for budget values ranging from CAN$5M to CAN$1B, in increments of CAN$5M. The results of these analyses are shown in Figures 2 and 3, which highlight how INMB and health benefit grow within each portfolio size as the adult immunization budget is increased. The cost-effectiveness approach reaches its maximum potential INMB for all portfolio sizes at a budget of roughly CAN$400M; the health-maximizing approach reaches its maximum health benefit for all portfolio sizes at a budget of roughly CAN$700M. For the full table of results at budgets of CAN$100M and CAN$1B see Appendix A (Table B.3 and Table B.4). Similar figures are available in Appendix B for the sensitivity analysis using the full cost of each vaccine, looking at solutions across a range of budget values under CAN$30,000/QALY (Figure B.5), CAN$50,000/QALY (Figure B.6), and CAN$100,000/QALY (Figure B.7) cost-effectiveness thresholds, as well as the health-maximizing approach (Figure B.8).

**Figure 2.**
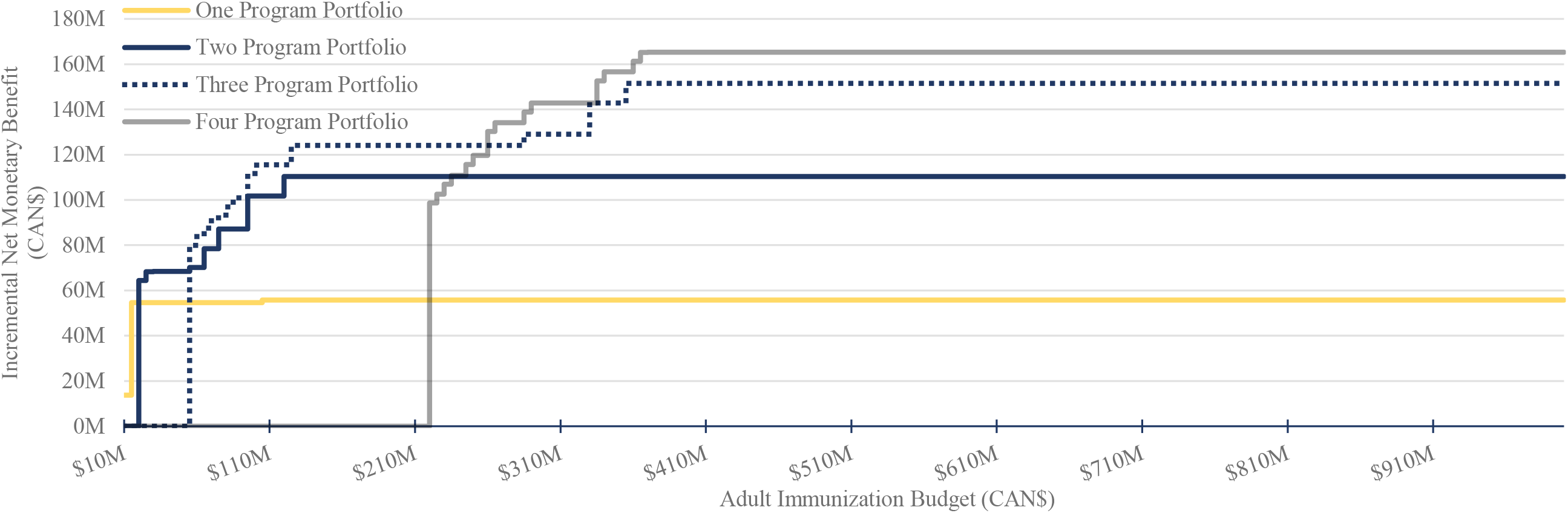
Main Scenario, Incremental Net Monetary Benefit Optimization at CAN$50,000/QALY

**Figure 3.**
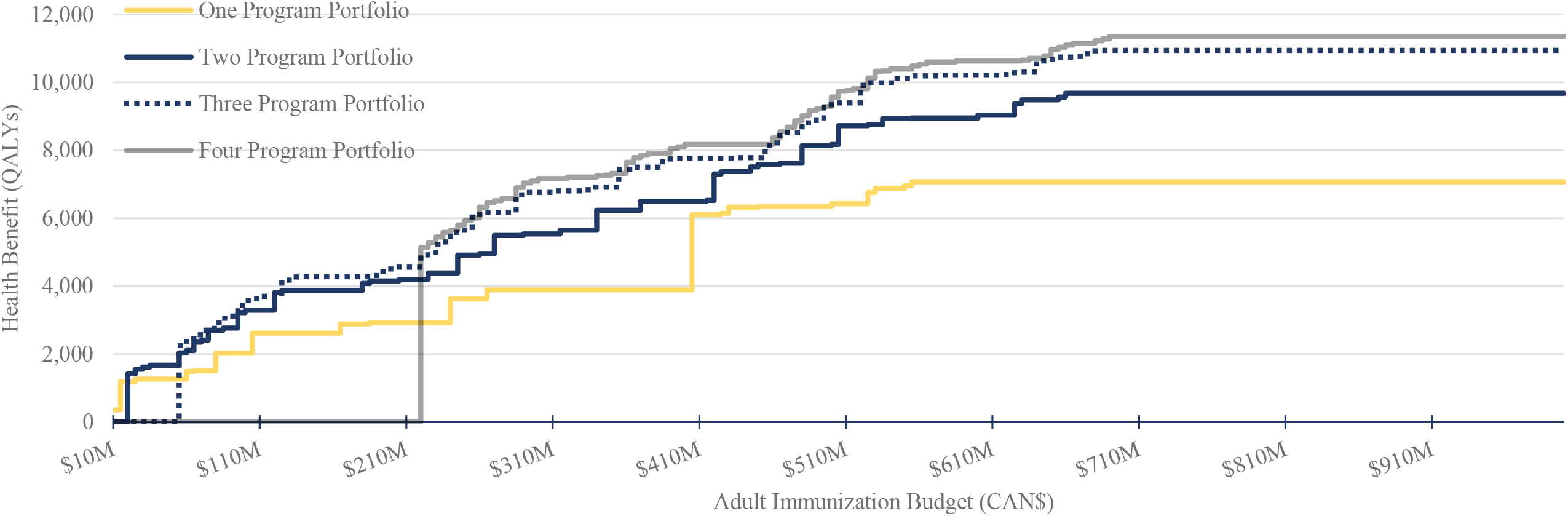
Main Scenario, Health-maximizing Optimization

## IV. Discussion

In this analysis, we optimized the adult immunization portfolio in Alberta based on INMB and health benefit. We found that the optimized solutions changed dramatically based on the number of immunization programs available, total budget, what we optimized for (i.e., health or INMB), the cost-effectiveness threshold and the assumed vaccine prices. When looking across the entire adult immunization program, we found that on average adult immunizations were not as good value for the money as what has previously been reported for childhood immunization programs (5), which are often found to be cost-saving (whereas none of the adult programs were found to be cost-saving). In comparison, at a CAN$30,000 cost-effectiveness threshold, even when assuming a 50% discount from the listed vaccine prices, there was no adult influenza program that was considered as part of the cost-effective optimization solution. Similarly, if we assumed no discount from vaccine list price at a threshold of CAN$30,000, only two programs were included in the final optimization model (Maternal Tdap [50% coverage] and Pneu-P-23 [at age 50]) using up only 5.67% of the CAN$500 million budget.

The chosen immunization programs, however, change substantially when optimizing health benefit instead of INMB at the CAN$30,000 threshold, with influenza vaccine programs having the highest health benefits, and therefore being the first program funded. This suggests that how budgets are distributed across public health and vaccines may have a big impact on which vaccine programs decision-makers selected for inclusion in the adult immunization portfolio. For instance, if budgets for the adult immunization portfolio are fixed, then the goal should be to maximize health benefits within that portfolio, with the potential to shift to more expensive programs that offer large health benefits. As observed in this analysis, a health maximizing approach will typically use up a higher percentage of the total budget, ensuring more health benefits for the population. However, if budgets can be spread across various programs, it may make more sense to optimize based on INMB, allowing budgets to be reallocated to more cost-effective programs within the public health or immunization portfolios (e.g., shifting more resources to improving coverage rates of childhood vaccines that are typically more cost-effective).

These findings demonstrate the complexity of decision-making around how to allocate scarce resources for immunizations and the need for methods and tools to allow these decisions to be made in real time with the best evidence available. By setting up an interactive optimization model, we allow for flexibility in decision-making, where we can incorporate new adult vaccines into the model as they become licensed or as immunization budgets change, providing an entire program perspective on where these new vaccines will fit within the larger adult immunization portfolio, and aiding in more efficient and transparent resource allocation decision-making.

While previous immunization studies have focused on constrained optimization of a specific vaccine program based on vaccines available (e.g., influenza, pneumococcal) or the efficient allocation of scarce vaccine resources (e.g., COVID-19 vaccines)(27-29), this is the first time to our knowledge of the approach being employed across an entire immunization portfolio. This study recognizes that a policy-decision based on one vaccine (e.g., to fund a new vaccine), can have big impacts on funding for other vaccines and health programs. By looking across the portfolio, the costs and benefits of multiple vaccination programs can be weighed simultaneously by policy-makers. As noted by Crown (2020)(30) and Standaert et al. (2020)(31), these methods are not being commonly applied within the healthcare system, but have the potential to support a range of programming decisions, including optimizing public health subject to a specific budget as we present here. Other areas of application include efficient delivery of supplies, health facility capacity management, staff scheduling, clinical decision-making, clinical trial design and optimal resource allocation, among others(32).

The analysis had a few limitations. First, since we estimated health outcomes associated with the different immunization programs using infectious disease models outside of the optimization model, we did not incorporate uncertainty around health outcomes in the optimization analysis. Future analyses could embed the infectious disease models directly in the optimization model. While this would allow for more complex analysis of uncertainty around the optimization experiment, this would substantially increase the run-time of the optimization model. Second, since the optimization results are based on the health outcomes estimated in the infectious disease models, they are subject to the limitations of each infectious disease model. While all the infectious disease models are Canadian-based and have undergone some level of peer review, they all include structural and methodological assumptions that may vary across the models and that should be taken into consideration when using the information produced by the optimization model. Third, there remains key uncertainties around vaccine price, which could substantially change the optimization results. In the main scenario we assumed a 50% reduction in listed vaccine prices; however, due to confidentiality surrounding government-negotiated vaccine prices, it is hard to judge whether this is a reasonable assumption. This highlights the importance of the interactive optimization model, where users can change input parameters, including vaccine price, and see how that impacts the optimization results. Fourth, we have only included vaccines that are publicly-funded by at least one province in Canada, and therefore vaccines funded through the federal government (e.g., COVID-19) and those coming down the pipeline at the time of our analysis (e.g., RSV vaccine for older adults) were not included in this initial analysis; however, these can be added to the future iterations of the analysis. Fifth, based on the population subgroups modelled in the infectious disease models, our ability to consider equity weighting for at-risk populations was limited. Through the interactive application, each immunization scenario can be assigned a specific cost-effectiveness threshold, which would allow weighting to be considered for at-risk groups were the necessary data to become available. Finally, Canada has no empirically derived cost-effectiveness threshold, which can substantially alter the vaccines selected as part of the cost-INMB optimization. Having an idea of the opportunity costs associated with resource allocation decisions would allow a more robust analysis of the tradeoffs associated with funding new vaccine programs.

This model could be expanded in several ways in the future. As noted above, the inclusion of vaccines in the pipeline for adults, as well as those currently funded through a different source, could help decision-makers who may need to make resource allocation decision about these vaccines in the near future. Moreover, more complex inclusion of uncertainty around health outcomes associated with each vaccination program could improve our understanding of optimization model unknowns. The methods used in this analysis could be included more broadly within the world of immunization, to evaluate the entire immunization portfolio, and provide decision makers a way to evaluate re-allocation of funds across a multitude of immunization programs. This more complete analysis would also have the added benefits of being able to effectively calculate a cost-effectiveness threshold for the immunization portfolio, which would also help with allocation decision-making. Future analyses could also add relevant constraints that were not included at this stage, such as vaccine administration constraints (e.g., the number of public health nurses or pharmacists available to vaccinate). Moreover, consideration could be given to how decision-making, and the factors considered, may change when adding a new vaccine to the portfolio versus optimizing an existing suite of immunization programs.

## V. Conclusion

This model demonstrates the informative power behind the constrained optimization approach when evaluating an entire portfolio of immunizations, rather than the current system of evaluating each new immunization program individually. As more adult vaccines come down the pipeline, it is essential decision-makers start to make allocation decisions within the larger context, especially as our analysis found adult vaccine programs may not be as good of value for the money, in comparison to childhood immunization programs or other public health interventions(4, 5).

Moreover, having optimization tools for a range of interventions available to decision-makers in real-time could improve goals-based decision-making, and improve effectiveness and transparency in making allocation decisions. This method requires decision-makers to think about the ultimate goals of their portfolios, along with any elements that may constrain their decision-making.

## Data Availability

The optimization model is available on request, and there is a publicly available version available online.

https://eshiny.ihe.ca/cvop/.

## Acknowledgements

We would like to acknowledge Danica Wolitski, Amy Elefson and staff at Alberta Health for the valuable expertise and feedback they shared with us. We would also like to thank Austin Nam for the use of the pneumococcal model he developed.

