## Supplemental File for "Optimization of an adult immunization program in Canada"

### APPENDIX A

**Table A.1** Coverage Rates, by Vaccination Grouping and Disease

| *Vaccination Grouping* | *Coverage Rate* | *Source(s)* |
|---|---|---|
| **Influenza** | | |
| Under 1 Year | 29.9 | (1) |
| 1-4 Years | 26 | |
| 5-9 Years | 17.2 | |
| 10-14 Years | 14.5 | |
| 15-19 Years | 12.7 | |
| 20-24 Years | 11.9 | |
| 25-29 Years | 14.4 | |
| 30-34 Years | 18.2 | |
| 35-39 Years | 20.2 | |
| 40-44 Years | 20.2 | |
| 45-49 Years | 21.5 | |
| 50-54 Years | 25.7 | |
| 55-59 Years | 31.6 | |
| 60-64 Years | 41.2 | |
| 65-69 Years | 53.7 | |
| 70-74 Years | 60.8 | |
| 75-79 Years | 66.1 | |
| 80-84 Years | 67.5 | |
| 85-89 Years | 70.5 | |
| 90+ Years | 74.5 | |
| Unknown | 0 | |
| All Ages | 26.8 | |
| 6 Months - 4 Years | 29.4 | |
| 65+ Years | 64.5 | |
| **Shingles** | | |
| Ages 70+ | 47.0% | (2-3) |
| Ages 65+ | 47.0% | |
| Ages 50+ | 27.4% | |
| **Pneumococcal** | | |
| Age 50 | 22.0% | (4) |
| Age 65 | 48.3% | |
| **Pertussis** | | |
| Pregnant women | 60.1% | (5-7) |

**Table A.2** Vaccine Effectiveness by Age, Any Influenza

| Product | aVE (65+) | aVE (20-64) | aVE (6-19) | aVE (<6) | Source |
|---|---|---|---|---|---|
| QIV-HD | 0.5105 | n/a | n/a | n/a | (8; Author Assumptions) |
| QIV | 0.3833 | 0.5100 | 0.5540 | 0.5540 | (9-11; Author Assumptions) |
| aQIV | 0.5309 | n/a | n/a | n/a | (12; Author Assumptions) |
| ccQIV | 0.4006 | 0.5336 | 0.5460 | 0.5460 | (9, 12; Author Assumptions) |
| aTIV | 0.4300 | n/a | n/a | 0.8600 | (13, 14; Author Assumptions) |

*Abbreviations:* QIV = quadrivalent; QIV-HD = quadrivalent high dose; aQIV = adjuvanted quadrivalent; aTIV = adjuvanted trivalent; ccQIV = cell-based quadrivalent; aVE = absolute vaccine effectiveness.

**Table A.3** Vaccine Effectiveness, by Age and Influenza Strain

| Product | aVE (65+) | aVE (20-64) | aVE (6-19) | aVE (<6) | Source |
|---|---|---|---|---|---|
| Influenza A | | | | | |
| QIV | H1N1 0.54<br>H3N2 0.33 | H1N1 0.55<br>H3N2 0.50 | 0.568 | 0.568 | (10, 11; Author Assumption) |
| ccQIV | n/a | n/a | 0.57 | 0.57 | (12; Author Assumption) |
| aTIV | H1N1 0.66<br>H3N2 0.40 | n/a | n/a | n/a | (Author Assumption) |
| Influenza B | | | | | |
| QIV | 0.31 | 0.45 | 0.495 | 0.495 | (Author Assumption) |
| ccQIV | n/a | n/a | 0.476 | 0.476 | (10, 11; Author Assumption) |
| aTIV | 0.285 | n/a | n/a | n/a | (Author Assumption) |

*Abbreviations:* QIV = quadrivalent; QIV-HD = quadrivalent high dose; aQIV = adjuvanted quadrivalent; aTIV = adjuvanted trivalent; ccQIV = cell-based quadrivalent; aVE = absolute vaccine effectiveness.

**Table A.4** Absolute Vaccine Effectiveness, Pneumococcal

| Product | Base | Source |
|---|---|---|
| Pneu-C (at age 65) | | |
| VT-IPD | 0.600 | (4) |
| ST3-IPD | 0.260 | |
| VT-CAP | 0.450 | |
| ST3-CAP | 0.156 | |
| Pneu-P-23 (at age 65) | | |
| VT-IPD | 0.470 | (4) |
| ST3-IPD | 0.020 | |
| VT-CAP | 0.200 | |
| ST3-CAP | 0.020 | |
| Vaccine effectiveness at age 50 | 1.1x effectiveness at age 65 | (4) |
| Vaccine effectiveness at age 75 | 0.9x effectiveness at age 65 | |

*Abbreviations:* VT = vaccine-type; IPD = invasive pneumococcal disease; ST3 = serotype 3; CAP = community-acquired pneumonia.

**Table A.5** Absolute Vaccine Effectiveness, Shingles

| Product | Ages 50-69 | Ages 70+ | Source |
|---|---|---|---|
| RZV | 0.97 | 0.91 | (16) |

RZV = recombinant zoster vaccine.

**Table A.6** Absolute Vaccine Effectiveness, Pertussis

| Product | Infants (Maternal vaccination) | Adult Booster | Source |
|---|---|---|---|
| Tdap | 0.80-0.90 | 0.8899 | (17-21) |
| | 0.80-0.90 | 0.8899 | |

Tdap = tetanus, diphtheria, & acellular pertussis.

**Table A.7** Full Vaccine Costs

| Product | Total cost, 1 dose | Source |
|---|---|---|
| Tdap | $78 | (22) |
| RZV | $165 | (22) |
| QIV | $46 | (22) |
| QIV-HD | $110 | (23) |
| aQIV | $113 | (23) |
| aTIV | $97 | (23) |
| ccQIV | $56 | (22) |
| Pneu-P-23 | $32 | (4, 24, 25; Author assumption) |
| Pneu-C-15 | $95 | (4, 24, 25; Author assumption) |
| Pneu-C-20 | $107 | (4, 24, 25; Author assumption) |

Abbreviations: QIV = quadrivalent; QIV-HD = quadrivalent High Dose; aQIV = adjuvanted quadrivalent; aTIV = adjuvanted trivalent; ccQIV = cell-based quadrivalent; Pneu-C-15 = 15-valent pneumococcal conjugate; Pneu-C-20 = 20-valent pneumococcal conjugate; Pneu-P-23 = pneumococcal polysaccharide 23-valent; RZV = recombinant zoster vaccine; Tdap = tetanus, diphtheria, & acellular pertussis.

4. National Advisory Committee on Immunization. Recommendations on the use of conjugate pneumococcal vaccine – 15 valent (PNEU-C-15) and 20 valent (PNEU-C-20) in adults:

Economic evidence supplementary appendix [Internet]. Date published: 2023-02-24. Accessed November 21, 2023 Available from https://www.canada.ca/en/public-health/services/immunization/national-advisory-committee-on-immunization-naci/public-health-level-recommendations-use-pneumococcal-vaccines-adults-including-use-15-valent-20-valent-conjugate-vaccines/economic-evidence-supplementary-appendix.html.

### APPENDIX B

**Table B.1** Vaccine Portfolio Selections, Full Vaccine Cost Scenario, CAN$500M Budget

| | | Size of Immunization Portfolio | | | |
|---|---|---|---|---|---|
| | | *One Program* | *Two Programs* | *Three Programs* | *Four Programs* |
| Health Benefit (QALYs Gained) | *HM* | 3,890.13 | 5,650.41 | 6,759.50 | 7,169.39 |
| | *INMB* | | | | |
| | *$50,000/QALY* | 1,193.89 | 1,412.68 | Not feasible | Not feasible |
| | *$30,000/QALY* | 1,193.89 | 1,412.68 | Not feasible | Not feasible |
| Percentage of Budget Used | *HM* | 83.06% | 99.78% | 93.67% | 97.50% |
| | *INMB* | | | | |
| | *$50,000/QALY* | 4.45% | 5.67% | Not feasible | Not feasible |
| | *$30,000/QALY* | 4.45% | 5.67% | Not feasible | Not feasible |
| Vaccine Program(s) Selected | *HM* | ccQIV for ages 18-64 | RZV for ages 65+ (57% coverage) **AND** QIV for 18-64 | Maternal Tdap (75% coverage) **AND** RZV for ages 50+ (37% coverage) **AND** QIV for 65+ | Maternal Tdap (75% coverage) **AND** RZV for ages 50+ (37% coverage) **AND** QIV for 65+ **AND** Pneu-C-20 & Pneu-P-23 at age 50 (22% coverage) |
| | *INMB* *$50,000/QALY* | Maternal Tdap (50% coverage) | Maternal Tdap (50% coverage) **AND** Pneu-P-23 at age 50 (22% coverage) | Not feasible | Not feasible |
| | *$30,000/QALY* | Maternal Tdap (50% coverage) | Maternal Tdap (50% coverage) **AND** Pneu-P-23 at age 50 (22% coverage) | Not feasible | Not feasible |

*Abbreviations:* HM, health-maximizing; INMB, incremental net monetary benefit; QALY, quality-adjusted life year; QIV = quadrivalent; ccQIV = cell-based quadrivalent; Pneu-C-20 = 20-valent pneumococcal conjugate; Pneu-P-23 = pneumococcal polysaccharide 23-valent; RZV = recombinant zoster vaccine; Tdap = tetanus, diphtheria, & acellular pertussis.

**Table B.2** Vaccine Portfolio Selections, Main Scenario, CAN$500M Budget and CAN$100,000/QALY Threshold

| | | Size of Immunization Portfolio | | | |
|---|---|---|---|---|---|
| | | *One Program* | *Two Programs* | *Three Programs* | *Four Programs* |
| Health Benefit (QALYs Gained) | *INMB* *$100,000/QALY* | 6,112.60 | 8,136.60 | 9,400.28 | 9,549.28 |
| Percentage of Budget Used | *INMB* *$100,000/QALY* | 80.39% | 95.70% | 99.74% | 99.25% |
| Vaccine Program(s) Selected | *INMB* *$100,000/QALY* | QIV for 18+ | RZV for ages 65+ (57% coverage) **AND** QIV for ages 18+ | Maternal Tdap (75% coverage) **AND** RZV for ages 65+ (57% coverage) **AND** QIV for ages 18+ | Maternal Tdap (50% coverage) **AND** RZV for ages 65+ (57% coverage) **AND** QIV for ages 18+ **AND** Pneu-P-23 at age 50 (22% coverage) |

*Abbreviations:* INMB, incremental net monetary benefit; QALY, quality-adjusted life year; QIV = quadrivalent; Pneu-P-23 = pneumococcal polysaccharide 23-valent; RZV = recombinant zoster vaccine; Tdap = tetanus, diphtheria, & acellular pertussis.

**Table B.3** Vaccine Portfolio Selections, Main Scenario, CAN$100M Budget

| | | Size of Immunization Portfolio | | | |
|---|---|---|---|---|---|
| | | *One Program* | *Two Programs* | *Three Programs* | *Four Programs* |
| Health Benefit (QALYs gained) | *HM* | 2,024.00 | 3,287.68 | 3,572.59 | Not feasible |
| | *INMB* | | | | |
| | *$50,000/QALY* | 1,193.89 | 3,217.89 | 3,572.59 | Not feasible |
| | *$30,000/QALY* | 1,193.89 | 3,217.89 | 3,572.59 | Not feasible |
| Percentage of Budget Used | *HM* | 76.55% | 96.79% | 98.64% | Not feasible |
| | *INMB* | | | | |
| | *$50,000/QALY* | 13.49% | 90.04% | 98.64% | Not feasible |
| | *$30,000/QALY* | 13.49% | 90.04% | 98.64% | Not feasible |
| Vaccine Program(s) Selected | *HM* | RZV for age 65+ (57% coverage) | Maternal Tdap (75% coverage) **AND** RZV for ages 65+ (57% coverage) | Maternal Tdap (50% coverage) **AND** RZV for ages 65+ (57% coverage) **AND** Pneu-C-20 at age 50 (22% coverage) | Not Feasible |
| | *INMB* | | | | |
| | *$50,000/QALY* | Maternal Tdap (50% coverage) | Maternal Tdap (50% coverage) **AND** RZV for ages 65+ (57% coverage) | Maternal Tdap (50% coverage) **AND** RZV for ages 65+ (57% coverage) **AND** Pneu-C-20 at age 50 (22% coverage) | Not Feasible |
| | *$30,000/QALY* | Maternal Tdap (50% coverage) | Maternal Tdap (50% coverage) **AND** RZV for ages 65+ (57% coverage) | Maternal Tdap (50% coverage) **AND** RZV for ages 65+ (57% coverage) **AND** Pneu-C-20 at age 50 (22% coverage) | Not Feasible |

*Abbreviations:* HM, health-maximizing; INMB, incremental net monetary benefit; QALY, quality-adjusted life year; Pneu-C-20 = 20-valent pneumococcal conjugate; RZV = recombinant zoster vaccine; Tdap = tetanus, diphtheria, & acellular pertussis.

**Table B.4** Vaccine Portfolio Selections, Main Scenario, CAN$1B Budget

| | | Size of Immunization Portfolio | | | |
|---|---|---|---|---|---|
| | | *One Program* | *Two Programs* | *Three Programs* | *Four Programs* |
| Health Benefit (QALYs Gained) | *HM* | 7,070.02 | 9,679.34 | 10,943.01 | 11,352.90 |
| | *INMB* | | | | |
| | *$50,000/QALY* | 2,609.32 | 3,803.21 | 7,429.62 | 7,839.51 |
| | *$30,000/QALY* | 1,193.89 | 3,217.89 | 3,572.59 | Not feasible |
| Percentage of Budget Used | *HM* | 55.42% | 65.68% | 67.70% | 68.90% |
| | *INMB* | | | | |
| | *$50,000/QALY* | 10.25% | 11.60% | 35.33% | 36.53% |
| | *$30,000/QALY* | 1.35% | 9.00% | 9.86% | Not feasible |
| Vaccine Program(s) Selected | *HM* | ccQIV for 18-64 and aQIV for 65+ | ccQIV for 18-64 and aQIV for 65+ **AND** RZV for ages 50+ (37% coverage) | Maternal Tdap (75% coverage) **AND** ccQIV for 18-64 and aQIV for 65+ **AND** RZV for ages 50+ (37% coverage) | Maternal Tdap (75% coverage) **AND** ccQIV for 18-64 and aQIV for 65+ **AND** RZV for ages 50+ (37% coverage) **AND** Pneu-C-20 & Pneu-P-23 at age 50 (22% coverage) |
| | *INMB* | | | | |
| | *$50,000/QALY* | RZV for ages 50+ (37% coverage) | Maternal Tdap (50% coverage) **AND** RZV for ages 50+ (37% coverage) | Maternal Tdap (50% coverage) **AND** QIV for ages 18-64 **AND** RZV for ages 50+ (37% coverage) | Maternal Tdap (50% coverage) **AND** QIV for ages 18-64 **AND** RZV for ages 50+ (37% coverage) **AND** Pneu-C-20 & Pneu-P-23 at age 50 (22% coverage) |
| | *$30,000/QALY* | Maternal Tdap (50% coverage) | Maternal Tdap (50% coverage) **AND** RZV for ages 65+ (57% coverage) | Maternal Tdap (50% coverage) **AND** RZV for ages 65+ (57% coverage) **AND** Pneu-C-20 at age 50 (22% coverage) | Not Feasible |

*Abbreviations:* HM, health-maximizing; INMB, incremental net monetary benefit; QALY, quality-adjusted life year; aQIV = adjuvanted quadrivalent; QIV = quadrivalent; ccQIV = cell-based quadrivalent; Pneu-C-20 = 20-valent pneumococcal conjugate; Pneu-P-23 = pneumococcal polysaccharide 23-valent; RZV = recombinant zoster vaccine; Tdap = tetanus, diphtheria, & acellular pertussis.

**Figure B.5** Scenario with Full Vaccine Costs and CAN$30,000/QALY Threshold, Incremental Net Monetary Benefit Optimization

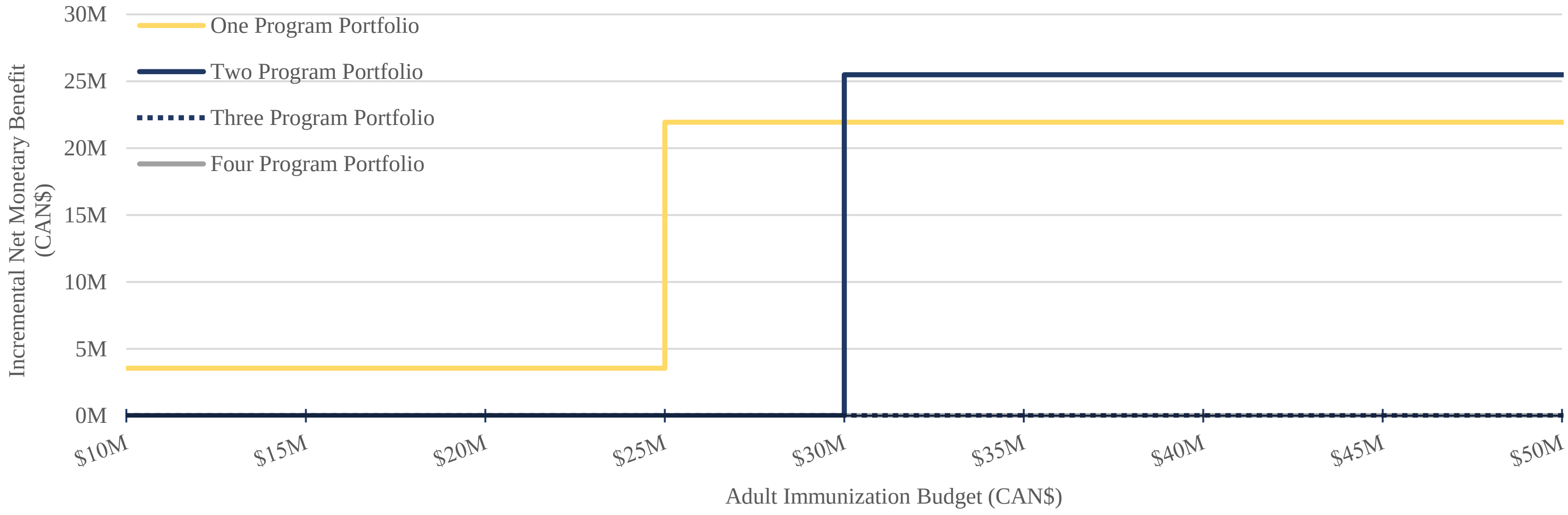


**Figure B.6** Scenario with Full Vaccine Costs and CAN$50,000/QALY Threshold, Incremental Net Monetary Benefit Optimization

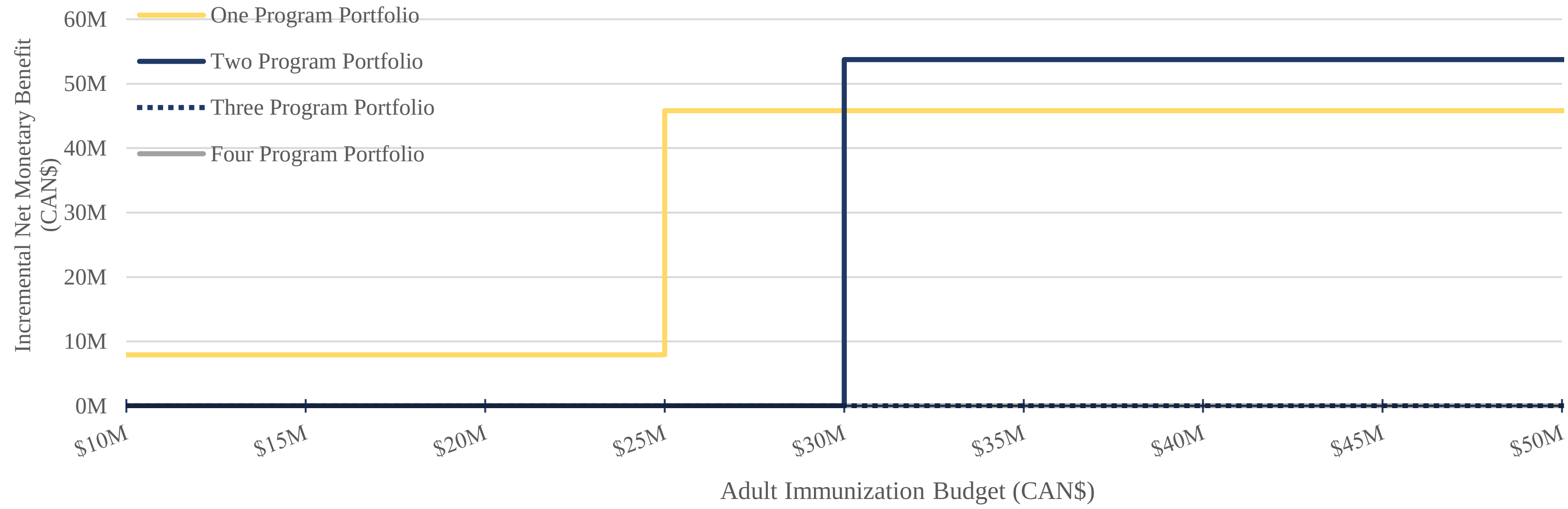

**Figure B.7** Scenario with Full Vaccine Costs and CAN$100,000/QALY Threshold, Incremental Net Monetary Benefit Optimization

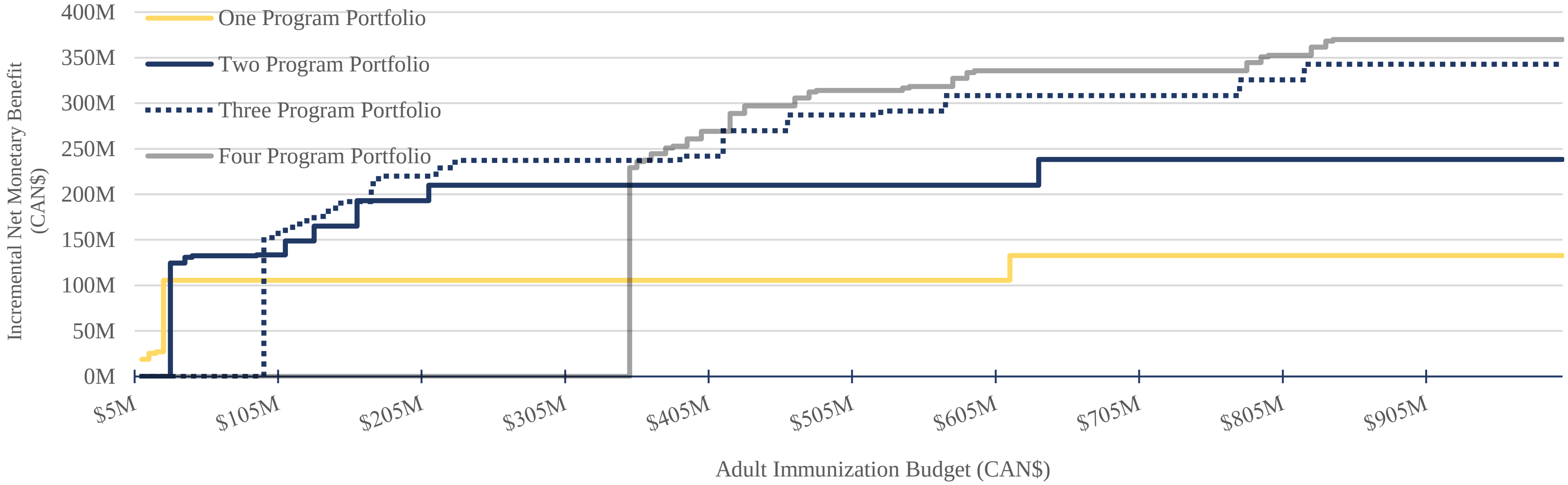


**Figure B.8** Scenario with Full Vaccine Costs, Health-maximizing Optimization

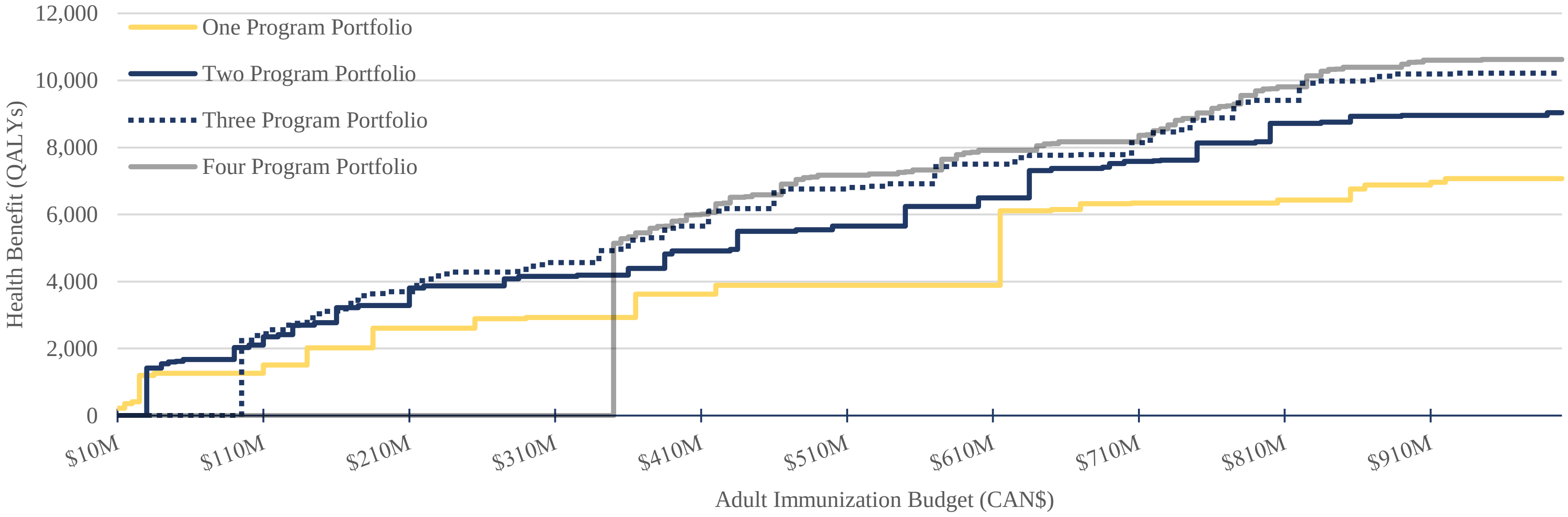
